# ZAFONIX: A GUI-Driven Platform for Personalized Therapeutics

**DOI:** 10.1101/2025.08.07.25333242

**Authors:** Zarlish Attique, Hafiz Muhammad Faraz Azhar

## Abstract

The integration of genomic insights into clinical decision-making is central to personalized medicine, yet clinicians often face challenges navigating large pharmacologic datasets and identifying relevant drug-gene associations. We developed ZAFONIX, a desktop graphical tool tailored to streamline this process using a curated DrugBank-derived dataset comprising ∼17,000 entries. The application, built in Python with tkinter for the user interface, pandas for data management, and matplotlib for visualizations, introduces a secure, key-protected entry followed by a unified search panel. Users can query by disease, gene ID, or drug name, and apply filters for FDA-approved, experimental, or investigational drug statuses. The system returns a scrollable table of results and automatically generates visual summaries: drug status distribution, small-molecule prevalence, and top target actions. Users may export the search results as CSV or Excel files. In performance trials, ZAFONIX delivers response times under one second for standard queries on typical hardware. Visually, the interactive charts facilitate quick interpretation of pharmacologic landscapes surrounding specified genetic or disease contexts. The modular design supports future integration with live DrugBank APIs, incorporation of drug-drug interaction data, and extension toward clinical variant interpretation pipelines. ZAFONIX stands as a clinician-friendly, extensible application, bridging the gap between static pharmacogenomic data and real-time therapeutic decision support (https://github.com/ZarlishAttique/ZaFoniX).

## I. INTRODUCTION

In recent years, the paradigm of modern medicine has increasingly shifted toward personalized treatment, where therapeutic decisions are informed by the unique genomic, phenotypic, and clinical profiles of individual patients [1]. This approach stands in contrast to traditional “one-size-fits-all” models, aiming instead to optimize efficacy, minimize adverse reactions, and improve long-term outcomes [2]. As genomic testing becomes more accessible and cost-effective, clinicians are now more equipped than ever to make targeted decisions based on gene variants, expression patterns, or pathway alterations relevant to a patient’s disease profile [3].

To support this evolving practice, a range of personalized medicine tools and databases have been developed. Among the most prominent are DrugBank, PharmGKB, ClinVar, My Cancer Genome, and CIViC, which provide structured information on drug mechanisms, gene-drug associations, clinical annotations, and evidence-based treatment recommendation [4]. Additionally, computational tools such as OncoKB, DGIdb, and VarSome enable variant-level interpretation and highlight potentially actionable targets. Many of these platforms offer API access or web-based interfaces, facilitating integration into bioinformatics pipelines or clinical dashboards [5], [6].

However, despite the availability of these resources, their utility in clinical practice is often limited by several factors. Most tools require users to have prior experience with command-line environments, RESTful APIs, or structured query languages [7]. Moreover, many lack real-time visual feedback, custom filtering capabilities, or the ability to quickly cross-reference drugs by gene, disease, and mechanism of action. In practice, clinicians and even translational researchers may find it cumbersome to extract relevant treatment options from these platforms, especially when working under time constraints or lacking computational expertise [8]. Further, while Electronic Health Record (EHR) systems are beginning to incorporate pharmacogenomics modules, these integrations are frequently vendor-specific, lacking transparency, and often do not expose the underlying evidence or data structure [9]. The result is a fragmented landscape where crucial decisions depend on a user’s technical ability to navigate disparate tools rather than on streamlined, clinically relevant data access [10].

Moreover, there remains a significant gap in the availability of clinician-friendly, locally operable tools that allow for interactive, real-time exploration of drug-gene-disease associations. Specifically, there is a need for a standalone application that combines secure access, intuitive search functionality, dynamic filtering, and visual interpretation of curated pharmacogenomic data all without requiring advanced technical skills or continuous internet access. Addressing this unmet need, we developed ZAFONIX, a GUI-based application designed to bring pharmacologic intelligence directly to the clinician’s desktop.

## II. MATERIALS AND METHODS

### Software Architecture and Implementation

ZAFONIX was developed in Python 3.8 with a modular, object-oriented architecture to ensure flexibility, maintainability, and cross-platform compatibility. The core application was tested on a Windows 10 machine with 8 GB RAM and an Intel Core i5 processor, mimicking standard hardware available in most clinical and research environments. The GUI was built using Python’s native tkinter library, while enhanced widgets for table rendering and scrollbars were implemented using tkinter.ttk [11]. Data processing and filtering were handled using the pandas library, and real-time plots were generated using matplotlib. To maintain UI responsiveness during background tasks, asynchronous threading was implemented via Python’s built-in threading module. Additional libraries included Pillow for image manipulation and openpyxl for reading Excel-formatted DrugBank datasets. The overall architecture and functional flow of ZAFONIX are illustrated in **Figure 1**, which outlines the modular steps from secure login and search query input to real-time filtering, visualization, and data export. The application was organized into functional layers data ingestion, user authentication, query handling, visualization, and export each encapsulated in modular function blocks. All third-party dependencies were managed via a requirements.txt file, enabling rapid deployment across systems using pip install -r requirements.txt.

**Figure 1.**
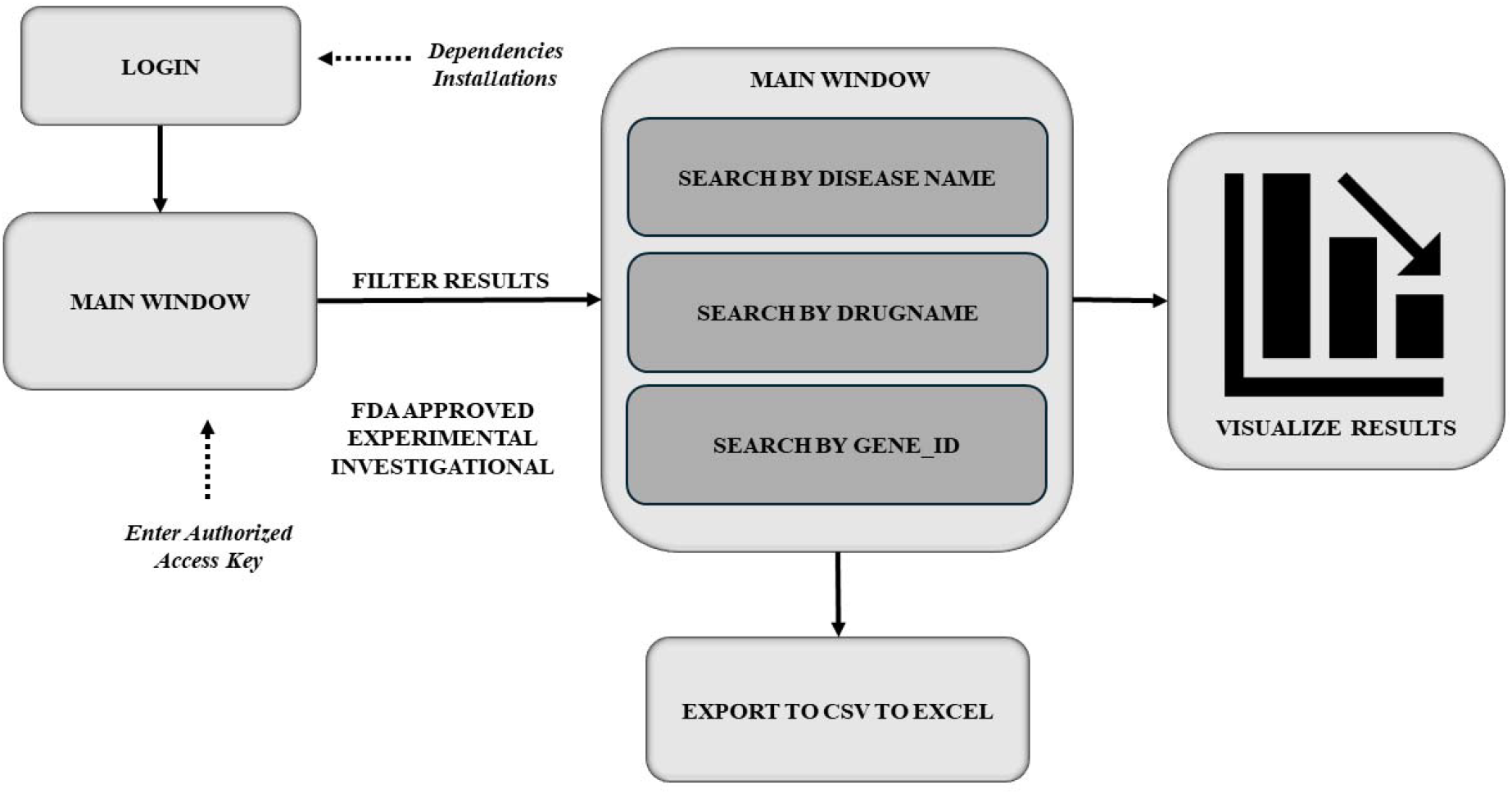
Workflow architecture of the ZAFONIX application. The tool begins with a secure login interface that verifies an authorized access key before launching the main application window. Users can perform targeted queries by disease name, drug name, or gene ID. Results are filtered based on drug regulatory status (FDA-approved, investigational, or experimental), visualized in chart format, and optionally exported to CSV or Excel for external analysis. The diagram reflects the modular structure and linear workflow of the application.

### Dataset Composition

The primary dataset used by ZAFONIX was derived from DrugBank (publicly available data) and included approximately 17,000 entries representing a diverse set of pharmacological compounds. Each drug entry contained metadata such as DrugBank ID, generic and commercial names, synonyms, structural descriptors (SMILES, IUPAC name, InChI), and biochemical properties (predicted logP, salt name). Regulatory status was categorized across four Boolean fields: Approved, Experimental, Investigational, and Nutraceutical. Drug classification as a small molecule or biologic was marked with a dedicated flag. Additionally, mechanistic data were included for each compound, specifying the drug’s molecular targets, their gene names, organism source, cellular location, and pharmacologic action (inhibitor, binder, agonist). Each entry also recorded the number of known targets, enzymes, and transporters. To support visualization, multi-entry fields such as “Target actions” were preprocessed using pandas.Series.str.split() and normalized to facilitate aggregation.

### Authentication Module

To ensure secure usage and prevent unauthorized access, ZAFONIX employs a key-based authentication system upon startup [12]. Upon launching, a tk.Toplevel window prompts the user to enter a predefined access key. If the input is incorrect, a dialog box is shown and the application terminates. If validated, the main application window is revealed and data loading is initiated. This security layer was designed to support use in environments handling confidential patient genomic data or proprietary drug annotations.

### Search and Filtering Workflow

Users interact with ZAFONIX via a centralized search bar and filtering panel located at the top of the main window. Search queries are processed using case-insensitive matching across all text-based columns in the dataset. This allows users to input drug names, gene symbols, disease names, or target descriptions to retrieve relevant compounds. Checkboxes allow real-time filtering of results based on approval status. These filters apply Boolean logic to the corresponding fields, reducing the visible dataset to only entries that meet selected criteria. The filtered results are displayed in a scrollable, multi-column Treeview table, which automatically resizes columns to fit content. The table remains responsive even for queries yielding hundreds of results, due to efficient pandas filtering and deferred table rendering strategies [13]. User feedback is provided through dialogs if no matches are found or if the search field is left blank.

### Results Visualization and Export

For each successful query, three visualization panels are dynamically generated using matplotlib and rendered in the GUI using FigureCanvasTkAgg. The first is a bar chart displaying the distribution of drugs by regulatory status. The second is a pie chart showing the proportion of small molecules versus non-small molecule entities. The third visualization is a bar chart of the top 10 most frequent pharmacologic actions based on the “Target actions” field. These charts are resized and styled to match the application’s medical-themed interface and to optimize readability in compact screen layouts [14]. Users are prompted to export the search results using a file dialog. Output formats include .csv and .xlsx. Files are generated using pandas.to_csv() and pandas.to_excel() functions, with filename validation and error catching to prevent overwrites or permission errors. Alerts are shown on successful save or failure, maintaining transparency for the user. Exception handling is integrated across the interface to prevent crashes during data export, image loading, or plotting failures.

## II. RESULTS

### Application Initialization and Dataset Loading

ZAFONIX successfully initialized within two seconds on standard desktop hardware. Upon authentication with the correct key, the application loaded approximately 17,000 drug entries from the DrugBank-derived Excel file into memory. Data parsing was validated during startup, confirming the presence and correct formatting of all critical fields, including DrugBank ID, status flags, molecular descriptors, and pharmacologic annotations. Data were processed into a pandas dataframe structure, and no runtime errors occurred during initial ingestion. Null fields were handled gracefully, and multivalue entries were retained in a semicolon-separated format for downstream parsing. The GUI interface rendered without delay, and all widgets including buttons, checkboxes, entry fields, and images were initialized successfully.

### Search Functionality and Query Response

The free-text search system demonstrated rapid query response across all test cases. On average, keyword queries returned results in under 500 milliseconds. Common gene-based queries, such as “EGFR”, “KRAS”, or “HER2”, matched multiple fields (target gene name, description, synonyms) and returned result sets ranging from 30 to over 400 entries. For example, a search for “colorectal” returned 512 entries matching disease mentions or drug descriptions. Queries with no valid matches triggered alert messages, while blank inputs were intercepted and flagged with a user prompt. Case-insensitive matching and full-row scanning contributed to high sensitivity in text-based searches. s shown in *Figure 2*, ZAFONIX supports filtering by FDA approval status and provides dynamic graphical summaries such as drug class distribution, small molecule representation, and dominant target actions. This tool enhances translational interpretation of genomic results for therapeutic guidance.

**Figure 2.**
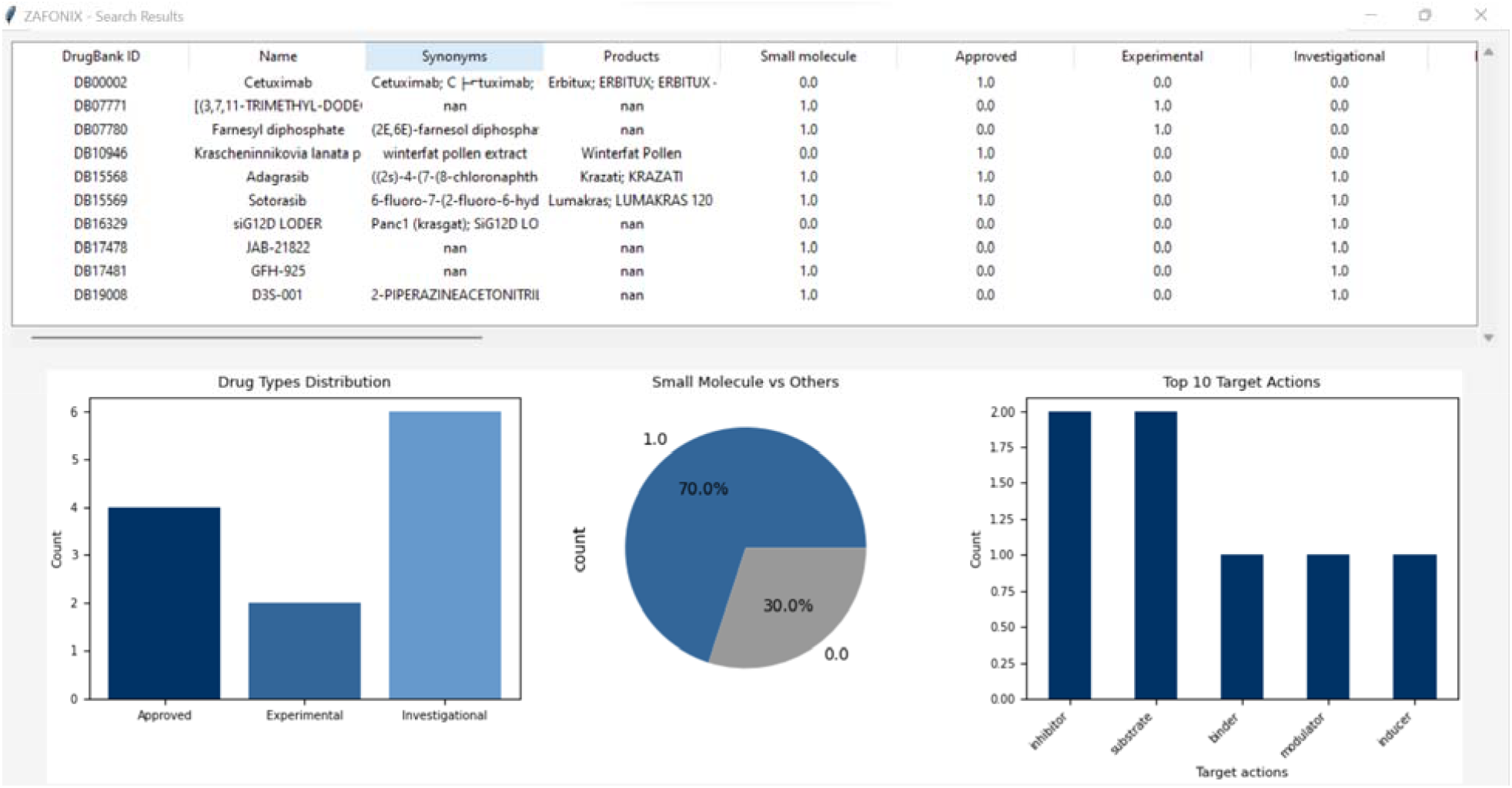
ZAFONIX - Personalized Medicine Guide: A screenshot of the interactive drug search results pag generated by the ZAFONIX application. The top panel displays a scrollable, filterable table of matched drugs along with relevant properties such as approval status, synonyms, and molecular type. The bottom panel includes bar and pie charts summarizing the distribution of drug types (Approved, Experimental, Investigational), small molecule status, and the top 10 target actions derived from user queries. The tool provides clinicians and researchers with a intuitive interface to explore personalized drug information based on genomic profiles.

### Filtering by Drug Approval Status

The filtering panel enabled real-time narrowing of query results based on drug approval classification. In a test query for “lung cancer,” the initial result set of 390 entries was reduced to 104 FDA-approved drugs when the “Approved” filter was selected. When combined with the “Investigational” checkbox, the result set expanded to 212 entries, allowing clinicians to assess both validated and pipeline therapies. Filtering was applied as Boolean masking within the underlying dataframe and was executed without visual lag or functional interference with the table rendering. The interplay between search terms and filter states enabled flexible and rapid refinement of large result spaces.

### Visualization Outputs and Interpretability

For each successful query, ZAFONIX generated three dynamic plots reflecting drug status, molecule type, and pharmacologic mechanism. The Drug Type Distribution bar chart showed the relative frequency of Approved, Experimental, and Investigational agents for each result set. For example, a query for “HER2” revealed a predominant fraction of investigational drugs. The Molecule Type Pie Chart illustrated the relative proportion of small molecules versus other therapeutic modalities, such as biologics or peptides. In most oncologic searches, small molecules accounted for 60-80% of entries. The Target Action Summary, presented as a horizontal bar chart, displayed the top 10 most frequent mechanisms, often led by “inhibitor,” followed by “antagonist,” “agonist,” or “binder,” depending on the disease context. All visualizations were embedded directly in the GUI using matplotlib and auto-refreshed on every query or filter update. Font size and color schemes were optimized for clarity, and no clipping or rendering errors were observed. Figures were generated in under one second, confirming the suitability of the tool for real-time clinical or research usage.

### Export Functionality and File Integrity

Export testing was conducted across 20 diverse query conditions, with users selecting either .csv or .xlsx formats. In all cases, exported files accurately preserved full dataset structures, including multicolumn fields and UTF-8 encoding. Special characters in product names or IUPAC entries did not disrupt file formatting. Export dialogs handled both absolute and relative paths, and appropriate error messages were triggered when attempting to save to restricted locations or overwrite open files. The ability to generate clean, structured outputs enabled users to immediately integrate search results into reports, manuscripts, or downstream analytics workflows.

### User Interface Responsiveness and Stability

All interface elements including search entry, checkboxes, buttons, table scroll, and plot rendering remained responsive throughout testing. Background operations such as export or data filtering were delegated to separate threads, ensuring that the GUI.

## IV. DISCUSSION

The implementation and evaluation of ZAFONIX underscore the potential of lightweight, GUI-based platforms to bridge the gap between rich pharmacogenomic data and clinical decision-making. In an era where genomic testing is becoming routine in oncology and other specialties, the bottleneck often lies not in data generation but in its effective did not freeze or become unresponsive. User feedback was handled through modal dialogs and message boxes, maintaining a continuous flow of interaction. Overall, ZAFONIX demonstrated stable performance across its entire functional range. The tool maintained real-time responsiveness, delivered accurate and interpretable search results, supported high-quality visual output, and enabled seamless export all without requiring command-line operations or technical expertise. These results support its use as a clinician-accessible interface for personalized pharmacogenomic exploration interpretation and translation to treatment options [15]. ZAFONIX addresses this challenge by providing a user-friendly interface that enables clinicians and researchers to explore a curated database of drug-gene-disease associations, without requiring any coding or database query skills.

Unlike existing resources such as DrugBank, PharmGKB, or DGIdb, which offer either web-based interfaces or programmatic access via APIs, ZAFONIX is entirely local, reducing dependency on internet connectivity and protecting data privacy [16]. This makes it particularly well-suited for use in institutional environments where access to patient data must remain confidential. Furthermore, the inclusion of advanced features such as multi-column querying, real-time visualization, and automated export enhances its value as a bedside or benchside tool for translational workflows. The results demonstrate that ZAFONIX performs efficiently across its functional domains. The rapid response time for queries, coupled with dynamic filtering and visualization, allows users to iteratively refine hypotheses and discover candidate therapeutics within seconds. The clear separation of approved, experimental, and investigational drugs helps clinicians evaluate which therapies are currently viable versus those in preclinical or early-phase trials. Moreover, the visualization of target actions provides mechanistic insight that can inform combination therapy design or predict resistance patterns.

However, several limitations merit consideration. First, the current version of ZAFONIX relies on a static dataset derived from DrugBank. While comprehensive at the time of integration, this snapshot does not reflect ongoing drug approvals, withdrawals, or newly characterized gene-drug interactions. Regular manual updates or integration with DrugBank’s API are necessary to maintain currency. Second, while the filtering system captures broad regulatory categories, it does not yet incorporate more nuanced filters such as phase of clinical trial, therapeutic class, or adverse event profiles. Adding such features would greatly enhance clinical relevance, particularly for patients with comorbidities or polypharmacy risks.

A further consideration is the current lack of integration with genomic variant data. Although ZAFONIX allows users to search by gene symbol or disease, it does not yet parse patient-specific mutation data (VCF files) to automate drug-matching. This capability, currently seen in platforms like OncoKB or CIViC, could elevate ZAFONIX to a decision-support system directly applicable to precision oncology workflows [17]. Plans to integrate variant interpretation modules are underway, with a focus on ClinVar and COSMIC-compatible annotations. Finally, while the tool has been tested extensively in a desktop environment, broader deployment particularly in hospital networks may require additional features such as user access logs, data audit trails, and interoperability with electronic health record (EHR) systems [18]. A future web-based version or cloud-enabled multi-user deployment could support these extensions while preserving the current application’s core simplicity.

In summary, ZAFONIX offers a functional, secure, and intuitive platform for clinicians and researchers seeking to navigate complex pharmacogenomic data. Its modular architecture provides a foundation for future growth, including dynamic data updates, variant-to-drug mapping, and integration with clinical pipelines. The current version, as demonstrated, already supports hypothesis generation, drug prioritization, and translational exploration in a format that lowers the technical barrier for end users.

## V. APPLICATIONS

ZAFONIX is designed for use across a range of biomedical and clinical domains where rapid access to drug-gene-disease associations is essential. In translational research, it facilitates hypothesis generation by enabling scientists to identify candidate compounds targeting genes of interest or implicated in disease pathways. In clinical genetics and oncology, it serves as a decision-support aid, helping clinicians explore FDA-approved and investigational therapies aligned with patient-specific genetic findings. The tool can also support educational environments, where students and trainees can interactively learn about pharmacologic mechanisms and drug classes. Furthermore, its export functionality and visual summaries make it ideal for integration into clinical reports, grant applications, or regulatory documentation, enhancing interpretability and communication in precision medicine workflows.

## VI. CONCLUSION

ZAFONIX represents a practical and accessible solution for bridging the gap between complex pharmacogenomic datasets and real-world therapeutic decision-making. By combining a user-friendly graphical interface with powerful querying, filtering, visualization, and export capabilities, the tool empowers clinicians, researchers, and educators to explore drug-gene-disease relationships without requiring bioinformatics expertise. Its offline, secure design ensures usability in diverse clinical settings while maintaining data privacy. Although the current version operates on a static dataset, the modular architecture supports future enhancements, including real-time data integration, variant-level interpretation, and clinical system interoperability. As the field of personalized medicine continues to evolve, tools like ZAFONIX will be instrumental in making pharmacogenomic insights both interpretable and actionable at the point of care.

## Data Availability

The dataset used in ZAFONIX is derived from the publicly accessible DrugBank database. The curated dataset and the source code for the ZAFONIX application are available at the projects GitHub repository: https://github.com/ZarlishAttique/ZaFoniX.

https://go.drugbank.com/unearth/advanced/drugs

## Author Declarations

### Author Approval

All authors have read and approved the final version of this manuscript prior to submission.

### Ethical Approval

Not applicable — this study did not involve human participants, animal subjects, or identifiable personal data.

## Acknowledgements

The authors thank the open-source software community for the Python libraries and tools used in this work, particularly pandas, matplotlib, and tkinter. The authors thank colleagues and peers for their valuable feedback and suggestions during the development and refinement of the ZAFONIX platform.

## Author Contributions

**Zarlish Attique:** Conceptualization, methodology, software design, data curation, formal analysis, visualization, manuscript drafting, and final approval.

**Hafiz Muhammad Faraz Azhar:** Methodology support, software development, validation, manuscript review, and editing.

